# Role of peer-tutors with dementia in Recovery College dementia courses: an ethnographic account

**DOI:** 10.1101/2025.07.24.25332075

**Authors:** Linda Birt, Melanie Handley, Juniper West, Jarin Alam, Fiona Poland, Esme Moniz-Cook, Emma Wolverson, Geoff Wong, Corinna Hackmann, Bonnie Teague, Rachael Litherland, Christopher Fox, DiSCOVERY study team

## Abstract

**Background and objectives:** Receiving a diagnosis of dementia impacts life plans and can lead to feelings of hopelessness and social disengagement. Post-diagnostic support can help people adjust to and assimilate a changing identity. Recovery Colleges in the UK offer a specific form of post-diagnostic peer-led support. This study aimed to identify *what works for whom, in what circumstances, and why*, within co-produced, peer-facilitated Recovery College dementia courses.

**Research design and methods:** Using ethnographic observations and interview data from the DiSCOVERY study, a realist evaluation of Recovery College dementia courses, we examined data to specify the activities of peer-tutors and the mechanisms which shaped outcomes for people with dementia.

**Results:** Five Recovery College dementia courses were observed across four NHS mental health services in England. Post-course interviews were undertaken with 13 tutors (3 peer-tutors with dementia) and 32 attendees (8 people with dementia). We found that through co-facilitation of recovery-focused content by peer-tutors who have well developed facilitation skills, attendees appeared to mediate self-stigma, manage emotional uncertainty and make meaningful social connections in ways which engendered hope for the future.

**Discussion and implications:** Identifying the activity between peer-tutors with dementia and course attendees foregrounds key strengths of this distinctive form of post-diagnostic support. Future work could evaluate longer term outcomes for people with dementia attending recovery courses, before potentially expanding this form of post-diagnostic support.

## BACKGROUND AND OBJECTIVES

Being diagnosed with dementia has a life-changing impact on a person and their family. People may need to adjust to and potentially assimilate a changed identity as a person with dementia (Birt et al., 2017). There is no cure and newly-licensed medicines have limited effect (Walsh et al., 2024). With numbers of people with dementia increasing globally (Wittenburg et al., 2019) and the reported rise in referral rates to memory services, the UK National Health Service (NHS) primary diagnostic route (NHS England, 2024), there is urgent need to provide sustainable post-diagnostic support. Sustainability in this context is defined as: *“… patient-centred, focused on recovery, self-monitoring and independent living, and actively reduces the need for intervention.*” (Jethwa et al., 2024 p5). The period immediately following diagnosis is critical in enabling people to maintain a non-stigmatised social ‘self’ (Birt et al., 2020) and to build personal resilience (Windle et al., 2023) for coping with the combined psychological, physical, social and emotional effects of this progressive and life limiting diagnosis (Watts et al., 2014).

Peer-led education offers ways for people to actively adapt and problem-solve to live positively with dementia (Bamford et al., 2021; Jopling, 2017). UK policy identifies key areas for improving the quality of post-diagnostic support, including shifting focus to ‘living well’ with long-term conditions. A national strategy promoted dementia peer-support and learning networks, specifying an objective to empower people with dementia to make their own choices in future planning (Department of Health, 2009). This vision contrasts with lived experience reports of post-diagnostic support disempowering individuals, defined as Prescribed Disengagement™ (Low et al., 2018). Dementia peer-support initiatives are commonly available world-wide, they are often community-led and as such, may provide support but will not necessarily specifically enable self-reflection, learning and empowerment. Finding ways for people with dementia to activate their personal strengths and build positive relationships is especially important as there are variable levels of statutory post-diagnostic support (Dooley et al., 2025).

### Peer support through Recovery Colleges

Recovery Colleges are an international initiative to help people assimilate mental health diagnoses on their own terms and learn ways to live positively with their symptoms (Leamy et al., 2011). The delivery of psychoeducation and adult learning is premised on co-production between people with lived experience and health professionals. Courses focus on personal recovery, informed by the ‘CHIME’ framework tenets: Connection, Hope and optimism, Identity, Meaning and purpose, and Empowerment (Leamy et al., 2011). Where and how Recovery College courses are provided differ, but in the UK, many align with and complement statutory mental health services (see Supplementary File 1). Importantly, the Recovery College model provides flexible access to psychoeducation, peer-support and knowledge sharing from lived experiences in the post-diagnosis period. Courses are generally open access enabling people with lived experience, family or friend supporters and healthcare staff to self-enrol without having to go through a referral system (Perkins et al., 2012).

### Recovery Colleges and dementia

Courses specifically designed by and for people with dementia and their supporters are being added to the Recovery College offer. Nonetheless such courses are not available in all NHS mental health services (Lowen et al., 2019) and when they are, staff involved in diagnostic services often do not routinely signpost newly-diagnosed people to Recovery Colleges (Wolverson et al., 2024). Where dementia courses are provided the co-production and co-delivery between staff and people with lived experience is considered important to personal recovery from a dementia diagnosis (Kenny et al., 2016; West et al., 2022). However there is limited empirical evaluation, including of the causal mechanisms underlying this. A realist review reporting on Recovery College dementia courses led to initial programme theories on aspects relating to co-producing courses (Handley et al., 2024). The review results emphasized the need for: co-production; tackling stigma; embedding of personal recovery principles; and the importance of providing practical and emotional support to ensure people adjusting to a diagnosis of dementia can access and benefit from the courses (Handley et al., 2024). However, the review found little evidence on the experiences of those who attended these dementia courses. A realist evaluation reports programme theories relating to setting up, advertising and delivering dementia courses (in review Birt et al., 2025).

## RESEARCH DESIGN AND METHODS

In the DiSCOVERY study we conducted a realist evaluation of English mental health service delivered Recovery College courses using a case study design (Birt et al., 2023). Realist evaluation is used to identify programme theories explaining how interventions are thought to work for various stakeholders by formulating evidence-based programme theories. These theories set out how relationships between the underlying mechanisms and contextual factors lead to different outcomes (Pawson & Tilley, 1997).

Important in realist approaches is a detailed understanding of context. Context may be shaped by organization policies and rules, but is also impacted by cultural concepts such a intersubjective beliefs, norms and values. Collecting data through a focused ethnography, using a case study design, enables examination of cultural factors at play (Jones-Hooker & Tyndall, 2023).

This paper draws on data from ethnographic observations and realist interviews with tutors and attendees who had dementia. However, given that courses took place in group settings, where interactions observed could be supportive or dismissive, we also draw on relevant data from family supporters and staff attendees to specifically examine and define key outcomes for people with dementia in co-producing, facilitating and attending Recovery College dementia courses.

### Study aim

This study aimed to define ‘*what works for whom, in what circumstances, and why*’, in Recovery College dementia courses. This paper has a specific focus on people with dementia either in a peer-tutor role or as an attendee.

### Ethical approval

Ethical approval was granted by Coventry & Warwickshire Research Ethics Committee (22/WM/0215). This work is part of the DiSCOVERY research programme (reference NIHR131676, 2022-2024). Full protocol details are available at Birt et al. (2023). A person with dementia from the DiSCOVERY Partners in Research group attended the ethics committee review meeting alongside researchers.

### Recruitment and sample

Recovery Colleges within English statutory mental health services offering dementia courses were approached. Inclusion criteria were: a) staff, people with dementia, family supporters either coproducing material and/or co facilitating courses; and b) course attendees - predominantly people with dementia and family supporters but also health and social care staff and members of a Recovery College. Purposive sampling enabled us to recruit courses from diverse geographical locations that had a variety of delivery styles. We sent study information to each site and held discussions before seeking consent from potential participants. In each case, staff and peer-tutors gave written consent to the course being observed. Recovery College staff sent study information to people enrolled on the course at least 48 hours before and course attendees were advised researchers would be present. Attendees’ consent was secured before the course started; all people with dementia were able to give informed consent. Tutors and attendees were informed of the opportunity to take part in a post-observation interview for which we obtained their additional consent. Recovery College staff and dementia service managers were also offered an interview.

### Data collection

#### Focused ethnography

Two of three female researchers (LB, MH JW) observed each course; all had extensive experience of research with people with dementia. Observations aligned with ethnographic methods using short, focused field visits (Wall, 2015). At times researchers joined in activities such as icebreakers, but mostly they sat amongst the attendees discreetly observing and recording activities. A formal template was not used but notes recorded the setting, actors, materials shared, who led discussions, tutors and attendees’ reactions to materials, chat between actors and changes in attendees demeaner or involvement. Following each session, researchers debriefed and reflected together.

#### Realist interviews

Tutors and attendees were offered individual interviews following the courses; either virtual or in-person. Topic guides (see Supplementary File 2) were developed with stakeholder advisory groups (Manzano, 2016) drawing on findings from the realist review (Handley et al., 2024). Interviews served the dual purpose of understanding individual experiences of Recovery College dementia courses and testing initial theories of what worked and why.

### Data analysis

Data were pseudonymized. Interviews were transcribed verbatim from digital recording and identifiers removed to maintain anonymity. Handwritten field notes were typed up and a single document agreed for analysis; reflective memos were maintained. Data were managed in NVivo (QSR International, 2020) and analyzed using realist logic (Pawson & Tilley, 1997). Coding was deductive (informed by the review’s initial programme theories); inductive (derived from the collected data); and retroductive (inferences about mechanisms based on interpretations of data about underlying causal processes). LB, JA and JW then independently revisited coded data on NVivo to develop Context-Mechanism-Outcome-configurations (CMOCs) which set out causal explanations of what works, for whom, when and how. Twenty CMOCs were grouped to the key practical concepts they represented (available for corresponding author).

An important step in realist evaluation is to consider formal (or substantive) theories which provide underpinning explanations for how mechanisms work in specific contexts to lead to outcomes. The conceptual personal-recovery CHIME framework (Leamy et al., 2011) and theories underpinning a person-centered approach to dementia (Downs & Lord, 2017) were identified and mapped using a collaborative tool to create a visual diagram (Miro, 2022). Research team meetings identified further conceptual models that explain the value of collaborative and interactive processes in Recovery College dementia courses. These included theories of hope (Dufault & Martocchi, 1985) and how people with dementia create meaning in everyday social interactions (Kitwood, 1997).

### Advisory groups

Developing results were structured into pre-reading discussion papers and slide sets by LB and JW, to enable systematic deliberations with the Partners in Research advisory group (people with dementia) and Staff stakeholder groups, and the wider research team. This iterative process of data analysis continued at regular intervals throughout the 18-month data collection period. Data was enriched with reflections from these stakeholders who gave consent to recording of meetings and use of illustrative quotes. Meeting transcripts were date-stamped, reviewed and advisory group statements validating or contesting researcher interpretations were highlighted. Specifically, the experiences of the Partners in Research group related to co-producing and attending similar ‘living with dementia’ courses providing triangulation of the empirical data set.

## RESULTS

### Sample

Five recovery-focused dementia courses were observed in four NHS Mental Health Trusts. Four courses were fully embedded within their Recovery Colleges. A fifth course, although originally developed within the Recovery College, was now being delivered as part of NHS post-diagnostic provision. Twenty-three people with dementia were observed and of those, 9 were interviewed across the 5 case studies (see Table 1 for sample details). To protect site and participant anonymity, we refer to participants only by their role: staff tutor, peer-tutor with dementia, peer-tutor family supporter; or attendee and their status: person with dementia, family supporter, staff or volunteer. There was representation of gender and age across the sample, but limited ethnic diversity. See next page for sample details

**Table 1.** Sample Characteristics.

|  | *Case study A* | *Case study B* | *Case study C* | *Case study D* |  |
| --- | --- | --- | --- | --- | --- |
| *Method of delivery* | In person<br>2.5 hours<br>3 sessions over 3 weeks | Online<br>1 hour<br>3 sessions over 3 weeks | In person<br>2.5 hours<br>single session | Online<br>2 hours<br>Single session | In person<br>2 hours single session |
| *Delivery team* | 2 staff tutors<br>1 peer-tutor with dementia<br>2 peer-tutor family supporters | 2 staff tutors<br>1 peer-tutor family supporter | 1 staff tutor<br>1 peer-tutor with dementia<br>1 peer-tutor family supporter | 1 staff tutor<br>1 peer-tutor with dementia | 1 staff tutor<br>1 peer-tutor with dementia |
| *Attendees* | N=9<br>4 spousal dyads<br>1 service manager | N=11<br>2 people with dementia<br>8 family carers<br>1 staff member | N=13<br>4 people with dementia<br>5 family/friend supporters<br>1 volunteer<br>3 support staff | N= 20<br>4 people with dementia<br>7 family supporters<br>8 staff<br>1 support staff | N= 33<br>5 people with dementia<br>9 family supporter<br>18 staff members<br>1 support staff |
| *Interviews* | N=10<br>2 staff tutors<br>1 peer-tutor with dementia<br>2 peer-tutor family supporters<br>2 attendees with dementia<br>2 attendee family supporters<br>1 attendee service manager | N=7<br>2 staff tutors<br>1 peer-tutor family supporter<br>3 attendee family supporters<br>1 attendee service manager | N=10<br>1 staff tutor<br>1 peer-tutor with dementia<br>1 peer-tutor family supporter<br>2 attendees with dementia<br>2 attendee family supporters<br>1 attendee volunteer<br>1 support staff<br>1 service manager | N=5<br>1 staff tutor<br>1 peer-tutor with dementia<br>2 attendee family supporters<br>1 attendee staff member | N=12<br>4 attendees with dementia<br>6 attendee family supporters<br>2 attendee staff member |

Three thematic areas) describe critical ‘stand out’ moments which identify how context shapes outcomes for attendees with dementia: 1) Empowering and enabling non-stigmatized identities, 2) Managing uncertainty for positive reframing; 3) Connecting together enabling hope. We present results by providing illustrative examples from observation data, supplemented with interview data, alongside interpretations of the context and mechanisms at play and the related outcomes.

### Findings

**Theme 1.**
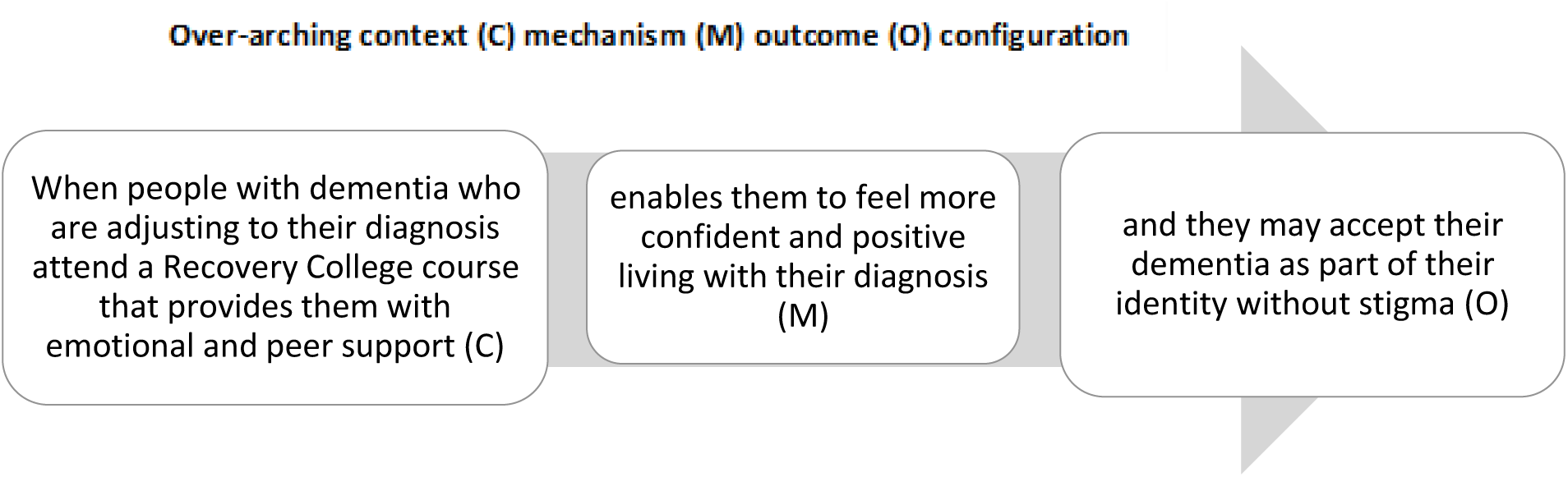
Empowering and enabling non-stigmatized identities.

An essential component in Recovery College courses is co-production and co-facilitation by peer-tutors with lived experience. The four courses where a peer-tutor with dementia actively co-facilitated, powerfully depicted how the actions of peer-tutors with dementia could be seen to enable attendees with dementia to experience positive connections, and importantly to also be seen by others as persons beyond their dementia designation, so helping to reduce self- and social stigma. Essential to inclusivity of course delivery was peer-tutors being seen as ‘one of the group’, more closely aligned with attendees with dementia than with staff. In some courses the peer-tutor clearly led the course, with the staff tutor merely prompting or helping to keep to time. In all in-person courses the peer-tutor at some stage positioned themselves physically with the attendees, and in one course the peer-tutor with dementia sat in the circle amongst attendees.

A ‘stand out’ moment occurred during an in-person, 2-hour single session course (Case D). The course was facilitated by a staff tutor and a peer-tutor with dementia; the peer-tutor predominantly led all activities with small prompts from the staff tutor. Attendees included staff, family supporters, and several dyads (person with dementia and family supporter). The peer-tutor recounted their traumatic experiences of diagnosis, and ways in which they now live their life more positively. This included how they maintain social connections, get involved in research and find purpose and meaning in everyday activities such as going for walks. Towards the session end, the staff tutor asks attendees to share experiences and for questions. Both tutors are sitting at the front of the room and chairs are set out in 5 rows with a central gap to walk down. Our observation fieldnotes capture ways in which the peer-tutor empowers an attendee with dementia to have a voice within a large and unfamiliar group of people.

> A person with dementia sitting in the back row next to his spouse starts to speak, he lifts a hand up, hesitantly stumbling to get words out… “*happy with you speech, can’t speak and I want to speak*” relating to what the peer-tutor with dementia is saying. The peer-tutor speaks directly to him and validates his experience; “*It’s frustrating, it is never easy*”. The attendee’s speech grows stronger as the peer-tutor nods towards him, and the attendee says, “*that’s exactly what I’m feeling, yes*”. The peer-tutor stands and moves forward into the gap between the rows of chairs to speak directly to the attendee, who then talks a bit about his previous occupation, “*used to work on a drawing board*”. His family supporter joins in to say the attendee might be embarrassed about his disabilities caused by dementia. The peer-tutor moves further forward and their one-to-one exchange continues, encouraged and reassured by the peer-tutor who says “*you put together the most wonderful sentence, don’t stop*” “*thank you for sharing*” and “*this is a very touching moment*”. Despite the direct interaction, the peer-tutor is simultaneously inclusive of everyone in the room by looking around and using physical presence to draw everyone in to the exchange, getting nods and murmurs of agreement from others. They conclude that people with dementia need more time to process and speak in each moment.

This extract demonstrates how important specific actions can be in promoting a non-stigmatized identity and /or felt sense of self for people with dementia. The actions that created the context (outlined in the overarching CMOC above) were the peer-tutor’s observation of the room and noticing a peer struggling to speak, their active positioning and ability to bring others into what was, in effect, a one-to-one interaction. The outcome was empowerment of the attendee with dementia including opportunity to position himself as a person by occupation, here describing his employment. A wider outcome was learning for others in the room about the importance of giving people with dementia time and attention to speak. On interviewing the peer-tutor with dementia afterwards, they reflected on this interaction: “… *but he put together this amazing sentence that just shows how, if people are in that environment where they feel safe, they can do more than they think they can. It just relaxes them into, for him, speech. … I mean, we have [standout moments] every time with peer support, but to have it happen with a stranger, it’s always just a magic moment*.” These may be examples of transformative learning (Oh, 2013).

By providing examples of living positively peer tutors appear to help attendees to have hope and consider themselves in more valued ways thereby helping to reduce self-stigma. The input from peer-tutors across all courses appeared to help family supporters and other attendees to consider people with dementia in non-stigmatizing ways.

In telling their experiences peer-tutors relived stigmatizing experiences and it was important they were empowered to have control over what they shared and when. Here in a post-session interview from Case A, the family supporter provides context to an event where the peer-tutor tells of sharing her diagnosis with a long-standing social group.

> ***Peer-tutor daughter***: Yeah, you do sometimes in some of the sessions, but not every session. So, I would say that when mum feels OK to share that she does and other times she doesn’t. (observation noted daughter looked to mother and checked she alright to tell story)
>
> ***Peer-tutor with dementia***: It’s a very difficult thing. They’d all just come back from a group outing and I’d got to tell them because I couldn’t just stand there because they were saying, “How are you?” I had to say, “I’ve got dementia.” There was everyone looking at everybody else and so in the end I just said, “Well we’ll go now then.”
>
> ***Interviewer to peer-tutor***: Yeah, so it sounds like each session, there is no script, you don’t always say the same thing, it’s whatever you feel is the right thing at that time.
>
> ***Peer-tutor with dementia***: Yes.

Here we see the family supporters work in creating safe spaces for people with dementia either as peer-tutors or attendees to revisit and share previous stigmatising events. When peer-tutors shared their diagnosis experience, attendees nodded as if agreeing and recognising their experience was not unique nor stigmatising.

**Theme 2.**
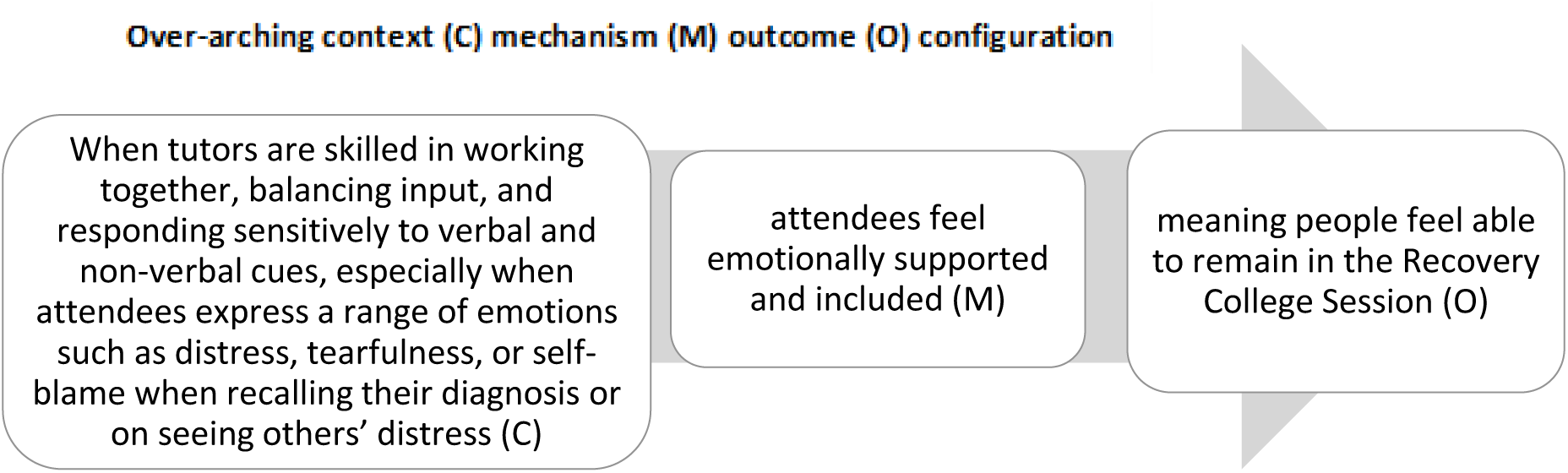
Managing uncertainty for positive reframing.

People usually attend Recovery College courses within a few months of diagnosis, meaning emotion and shock from receiving their diagnosis can still be raw. This can lead to people becoming distressed during the course. While staff tutors will usually be able to draw on clinical training to support an attendee who is emotionally upset, peer-tutors often do not have formal training for this. This ‘stand-out’ moment illustrates both the empathy peer-tutors can display with others’ lived experiences, and the skills they can adopt to contain upsetting emotions in others.

The course was in-person and a single 2.5-hour session (Case C). The setting was a meeting room in a building used by the mental health service. The course was facilitated by a staff tutor and two peer-tutors: a person with dementia and a family supporter There were 10 attendees: people with dementia attending alone, family supporters attending alone, 2 dyads (person with dementia and family or friend supporter) and a staff attendee. Attendees heard about the course mainly from their clinician, but a couple had heard from other community groups including the attendee who was distressed. Before the course started a person with dementia, attending alone, was visibly distressed, attendees waited with some hesitant anticipation for the course to start, a few people talking to each other. The extract below describes the situation approximately 10 minutes into the course; the group had introduced themselves by stating they had dementia or were a family or friend supporter.

> Everyone is sitting around a large central table which means people can have eye contact. The attendee shows small signs of anxiety through worried facial expression. She then becomes distressed and tearful, self-blaming for having dementia. Everyone stops and listens and the peer-tutor with dementia responds and offers reassurances. Following the exchange, the staff tutor takes a seat near to the computer for presenting slides and indicates non-verbally to the support staff member to move to the empty chair next to the person. A group activity is facilitated where peer-tutors hand round a wellbeing questionnaire for attendees to chat through together and complete. There are high levels of engagement by all with ‘ad hoc’ questions being asked about dementia, and some laughing and joking. The attendee again becomes tearful while talking of a traumatic experience, the peer-tutor with dementia takes over from the staff member sitting next to the person. They speak quietly and supportively and the attendee visibly calms. Following slides about the ageing brain, the staff tutor sets the scene for a further interactive activity, by asking “*What is your understanding of what it is like to have dementia? What images come to mind?*” The peer-tutor with dementia gets up; *“Let’s write some of these down”* and moves to a flipchart. The attendee again becomes distressed *“It’s my fault I’ve got it [dementia]. I blame myself”*. The staff tutor asks the person and the group to focus on dementia symptoms *“what sorts of symptoms do you have?”.* Attendees including the distressed attendee offer words such as ‘forgetfulness’ and the peer-tutor family support writes them on the flipchart, the peer-tutor with dementia talks about his own experience to keep the group joining in.

This example provides evidence of both the staff and peer-tutor being aware of others’ emotions and having skills and being prepared to work together to manage emotions. For example, here the staff tutor indicates for someone to sit next to the attendee and shifts the focus onto symptoms rather than causes. The peer-tutor with dementia showed awareness of the attendees’ struggle; and had the confidence to support with quiet interaction. The attendee is able to contain their distress, and the rest of the group can relax. The mechanism found to help resolve this situation appears to be the ability of tutors to create a space of psychological security, where a range of emotions can be acknowledged rather than dismissed. In the post-course interview, we spoke to the peer-tutor with dementia about the attendee’s distress and how the situation was managed. The peer-tutor explained they helped the attendee contain their emotion: “*I was worried. When the lady who presented with the very emotional, tearful response at the beginning, you know, she was on the verge of it being appropriate to remove her from there to somewhere quiet for a one-to-one chat*.” They go on to explain that if the staff tutor left the room this would leave the peer-tutors uncertain about what to do: “*I don’t know how I would’ve coped with that because the only person at the moment who is competent to deliver the rest of that presentation is the practitioner [staff tutor] who would probably have wanted to be the one who dealt with that lady*.

This highlights a challenge for delivering courses if there is reliance on only one or two people having the knowledge or skills to facilitate. Finally, the peer-tutor confides that the situation felt like a ‘*small pressure cooker room’* where ‘*hysteria could have taken over in the room*’. Importantly because all tutors were able to effectively help the attendee ‘hold’ their emotion, the person was calm enough to attend to the information being shared by the staff tutor on the potential causes of dementia, and later she is heard to say ‘*I don’t need to blame myself’*.

Further into the session the peer-tutor with dementia leads a section creating opportunities for the attendee with dementia to share positive aspects of herself beyond dementia.

> The staff tutor asks the peer-tutor with dementia to discuss their experiences, he engages the group, they are all listening. When speaking of their fundraising the previously distressed attendee with dementia chips in that she joined the fundraising walk. The peer-tutor passes pictures round of things they have done, there is some laughter as they talk. The peer-tutor describes being able to use ‘dementia goggles’; *“You are valuable as consultants”*. The attendee with dementia says ‘*you feel wanted’* there is a visible change in her facial expression and body language appears more upright and smiling, ‘*you meet people who make you laugh’*. The family supporter peer-tutor is writing down ideas on flipchart of things people suggest for wellbeing and the attendee with dementia says she does knitting and puzzles to help her mind.

This stand-out moment demonstrates the value of having peer-tutors with lived experience of dementia who have empathy with emotional responses, but also demonstrate ways in which they have managed the uncertainty of dementia themselves.

> The staff tutor asks the peer-tutor with dementia to discuss their experiences, he engages the group, they are all listening. When speaking of their fundraising and the previously distressed attendee with dementia chips in that she joined the fundraising walk. The peer-tutor passes pictures round of things they have done, there is some laughter as they talk. The peer-tutor describes being able to use ‘dementia goggles’; *“You are valuable as consultants”*. The attendee with dementia says ‘*you feel wanted’* there is a visible change in her facial expression and body language appears more upright and smiling, ‘*you meet people who make you laugh’*. The family supporter peer-tutor is writing down ideas on flipchart of things people suggest for wellbeing and the attendee with dementia says she does knitting and puzzles to help her mind.

This stand-out moment demonstrates the value of having peer-tutors with lived experience of dementia who have empathy with emotional responses but also demonstrate ways in which they have managed the uncertainty of dementia themselves.

**Theme 3.**
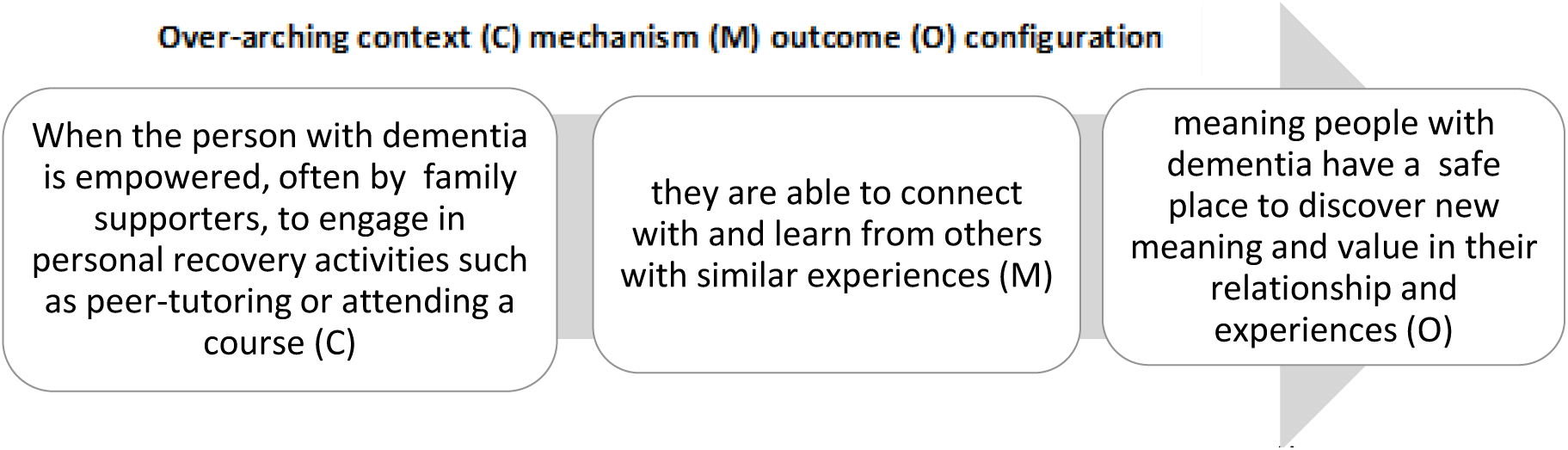
Connecting together enabling hope.

While a person with dementia may attend a Recovery College course alone it was more usual for people to attend as a dyad with a spouse, adult child or friend supporter. During in-person course we observed ways in which peer-tutors created content and time for dyads to undertake activities together, prompting connections in a ‘safe place’ away from usual home life. Having established this, the course moved onto several couple-based activities including one where each dyad discussed between themselves what made them feel stressed, again creating a space for the person with dementia to identify issues that increased stress. We noted corresponding phrases such as “*when you rush me to go out I feel stressed and anxious*”. We observed where activities asked dyads to work together, staff and peer-tutors often moved to be physically present with the dyad and to support conversations between individuals. Sometimes the person with dementia acting with openness in the safe setting of the Recovery College course deepened the connectedness between relatives. In this stand-out moment during an in person single 2.5-hour course (Case C), the relationship history between the person with dementia and the adult-child was difficult; the family supporter considered their parent to be in denial of their diagnosis, as they never spoken about having dementia.

> Once everyone had settled, the staff tutor stood at the front of the room, welcomed everyone to the session and invited round table introductions. Everyone used the word ‘dementia’ during their introduction although there was no stated prerequisite for this. Each person said why they were there, and the mother of one child/parent dyad clearly said three words “*I have dementia*”, at which the daughter showed complete surprise.

The daughter later explained during an interview how this incident enabled her to understand more of her parent’s experience and to begin to know how her parent felt about their diagnosis: *“It was the first time my mum had actually said that she had dementia there. … She’d not said it before, no. … That was the first time and she actually said those words and I was like, ‘Oh God, I can’t believe it.’ … I was really surprised and it made me feel quite emotional. You know, I could have easily burst into tears I think when she said it. … and it probably made me feel more connected to her because it’s difficult to be close to my mum”*.

This powerfully exemplified how a non-judgmental space within the Recovery College course enabled a person with dementia to verbally acknowledge this in front of their relative. Following this event when attendees were asked to complete a wellbeing questionnaire, we saw this couple talking together as the daughter helped her mother read and complete her form. All Recovery College courses promote an expectation that content will be person-centered, materials will draw on social models of health, common ‘clinician-patient’ power imbalances will be levelled, and people will connect together. However, in some courses we observed delivery of material being tutor-led with limited space or activities for attendees to connect. Factors like the setting and method of course delivery affected whether people had the time and physical space to connect informally or informally. For example, being in a warm and welcoming setting with space for coffee breaks and movement meant people interacted more. Here a peer-tutor with dementia who delivered courses both online and in person (Case D) explains the importance of such breaks for socially connecting and information sharing:

> Peer-tutor with dementia: *Also it may be a bit of a bizarre thing but you’d miss the coffee breaks [online] and we have really good conversations in coffee breaks. We will find out, we’ll talk to people and people are open and much more likely to say something that’s concerning them or something they might think it’s silly and don’t want to say in front of anyone else. All sorts of things have come out of previous groups in the coffee break haven’t they. We’ll talk about all sorts with people and you’d miss that*.

In courses run in-person across several sessions we observed interactions between attendees and peer-tutors increasing over time. Coffee breaks and food sharing led to an attendee with dementia striking up conversation and sharing recipes with another attendee. The person with dementia was thus enabled to present themself as a cake-baker and to discover shared interests with a person who was a stranger, socially connecting and empowering conversations beyond having a dementia diagnosis.

Person-centered activities offered opportunities for attendees with dementia to socially connect with others. We saw activities creating safe spaces for people with dementia to join in without fear of failure or need to explain their dementia. In warm up activities prefaced with ‘*tell us why you are here?’* people always stated their dementia status or supporter, or staff role. This created a status divide. In one course the peer-tutor suggested a more inclusive warm-up: people were asked if they were a dog or cat person. The activity was introduced using slides with images of cats and dogs and the peer-tutor stated their preference. Every attendee was able to contribute, there was laughter, talk of pets from childhood and when their families had been at home; this prompted shared accounts between spousal and child/parent dyads. If people liked neither cats nor dogs the staff tutor asked about any animal they did like. This led to an attendee with dementia hesitantly telling the group about the birds which they chased with sticks in the garden. They had difficulty with word-finding, but was given time to tell their account. As the group discussed this image and shared laughter, we observed the person sitting a little taller in their chair and looking around the room more actively. The staff tutor started each week reminding people of the previous week’s animal discussion, scaffolding more talk around this non-threatening topic. Both these examples demonstrate dementia course activities creating spaces for people to feel included and inter-connected to others.

## DISCUSSION AND IMPLICATIONS

### Summary of findings and comparison with existing literature

This is the first in-depth study to understand what might work for who and in what circumstances in Recovery College courses for people with dementia following a dementia-diagnosis. The concept of recovery-focused care being appropriate for people with dementia was raised as early as 2010 by Hill. However, ours seems to be the first empirical evaluation to identify the specific contextual and constituent features of Recovery College dementia courses and what outcomes can be for people with dementia in either peer-tutor or attendee roles. We used realist approaches to examine underlying causal processes, or the mechanisms, in Recovery College dementia courses. By developing middle-range theories, we aimed to understand what is happening in this complex intervention. Context, mechanism, outcome configurations (CMOCs) provided causal explanations from observation and interview data. The mechanisms set out within these CMOCs and related exemplars can be seen to illustrate theories which align with the CHIME framework, namely: Connectedness, Hope through positive reframing, non-stigmatized Identity and Empowerment, related here to dementia literature and post-diagnostic support principles.

#### Connectedness

Attending a Recovery College course can enable people with dementia to maintain a social self. By connecting and taking time to inter-relate with others with similar experiences, loneliness in people with dementia can be counteracted. Structuring the course with time for informal talk between attendees and tutors, connects people through shared interests or humor, not just through the dementia label. The importance of enabling different relationships beyond ‘person with lived experience’ and ‘family supporter’ has been highlighted in general Recovery College course evaluations (Reid et al., 2020; Toney et al., 2018) and in dementia literature (Birt et al., 2020). Additionally, as co-produced psychoeducation is a collaborative and interactive process that creates value through meaningful exchanges, attendees with dementia can connect despite differing levels of engagement, by interacting verbally or non-verbally, watching and listening. This resonates with Kitwood’s (1997) finding that positive social psychology upholds psychological needs in dementia such as inclusion, attachment and identity.

#### Hope through positive reframing

Meeting other people with dementia can help individuals reframe their experiences and foster hope. Generating and maintaining hope is an interpersonal process (Dufault & Martocchio, 1985; Miller, 1985) and after a dementia diagnosis, people may face challenges in adjusting expectations (Wolverson et al., 2010). Comparing their situation with others and learning coping strategies from peers can foster hope (Amponsem et al., 2023). Social interactions create opportunities for experiencing fun and to affirm people’s continuing capacity for positive experiences. Such reciprocity and reaffirming support help generate more hopeful experiences (Jackman et al., 2024).

#### Non-stigmatized identity

Our findings highlight the importance of creating and exemplifying non-judgmental spaces, non-stigmatizing spaces, where the contributions of people with dementia are of equal value to those of healthcare staff and family supporters. By encouraging, sharing and building peer experiences, people with dementia can recover a non-stigmatized life after diagnosis (Mukadam & Livingston, 2012). Peer-tutors with dementia bring authenticity to their interactions with attendees, where they can connect through sharing experiences in an immediate, validating, and empathetic way. Peer-tutors with dementia experienced positive outcomes such as developing their sense of self-worth and confidence. They were supported to accomplish complex occupational and social activities and could see the benefits that other attendees gained. These findings align with other evidence that people with dementia can and do maintain social connections and are involved in helping others, so sustain a non-stigmatized identity (Birt et al., 2020); affirming self-hood (Hennelly et al., 2021). A Recovery College dementia course is not only a social setting but also one which offers learning about dementia and experiences of how people can navigate life positively with dementia, so replacing guilt and shame with validation and empowerment. However, Reid and colleagues’ (2020) evaluation of a recovery education centre noted that deconstructing self-stigma was an ongoing practice. This highlighted for us that while peer-tutors in our study did demonstrate and articulated self-esteem and purposeful activity, this could not reliably evidence longer-term positive outcomes for attendees. Our CMOC (Theme One) indicated an outcome for people with dementia attending Recovery Colleges was to develop or maintain a stigma-free identity. Further evidence is needed on the longer-term impact of single courses and if longer-term sustainable post-diagnostics recovery focused support is needed (Moniz-Cook & Mountain, 2023).

#### Empowerment through safe uncertainty

People often attend Recovery College dementia courses quite soon after diagnosis. This can be a time when people feel emotionally unsafe as they face a health threat, and individuals and family supporters experience concerns about each other’s wellbeing (Moniz-Cook et al., 2006). The learning embedded in Recovery College courses may be essential to help people with dementia gain a sense of emotional safety (Kuske et al., 2021). Recovery-focused and person-centered post-diagnostic support gives control to the person with dementia. The ethos of encouraging people to be empowered and have choice even if these are deemed risky by others (Clarke et al., 2011), may be seen to reflect the theory of ‘safe uncertainty’ articulated by Mason, 2022. When under stress, people may wish for certainty and clear decisions made for them, but that this in itself may bring negative outcomes (Mason et al., 2022). A more therapeutic framing of options is to accept there are no definitive solutions or ways to sort the ‘problem’ (here a dementia-diagnosis}. Instead there needs to be a shared process of learning to manage the uncertainty that life brings and to use personal resources to deal with everyday life (Mason et al., 2022). Such active learning aligns with framing recovery as an ongoing and personal process (Leamy et al., 2011) as Recovery Colleges may empower people with skills to consider ways to manage future challenges (Hill et al., 2010). Having choice, control and self-determination are all associated with empowerment in people with dementia (van Corven et al., 2021). Mental health experience-based co-production initiatives are designed and shown to support people to make sense of real-life problems and find ways to move forward by learning from the past to shape the future, even when they are uncertain (Palmer et al., 2019).

### Study Limitations

We acknowledge the limitation of recruiting from just four sites where our case study ethnographic approach allowed in-depth observations of diverse course-delivery styles. Recovery College dementia courses are relatively new and require future research to evaluate the longer-term impact of this form of post-diagnostic support, and the impacts for different people. We sought, but did not achieve, an ethnically diverse sample despite recruiting from Recovery Colleges in areas with ethnically diverse communities. Known barriers to attending mental health services for both diagnosis and support might explain limited recruitment of non-white British people in this small sample. Despite our study being situated only in England our realist findings may be internationally transferable as Recovery Colleges run in over 28 countries (Hayes et al., 2023).

### Conclusion and recommendations

This study is the first to examine co-production, co-facilitated peer support within the context of Recovery College dementia courses - a distinct and novel form of recovery-focused post-diagnostic dementia support. In-depth case study methodology has offered themes associated with positive constructs such as connectedness, hope, empowerment and counteracting a stigmatised identity associated with a dementia-diagnosis. This study reflects a next step in identifying *what might work for who and in what circumstances*, following a dementia-diagnosis. We suggest the context mechanism and outcomes outlines in this paper set parameters for future research to design conceptually-driven post-diagnostic psychosocial intervention studies in dementia.

## Acknowledgements

The authors thank all those who took part in the study enabling us to observe sessions and speak with tutors and attendees across England. Thank you to the study manager Thomas Rhodes for expert guidance on protocol and ethical management. This study was actively supported by two advisory groups; we thank the Partners in Research (Patient and Public Involvement and Engagement) group who, with the support and expertise of Rachael Litherland from Innovations in Dementia, actively contributed to our analytical processes, providing perspectives from people living with dementia, and from family supporters. Thank you to all the NHS memory and dementia service and Recovery College staff who formed our staff advisory group helping to ensure outcomes resonated with practice needs. Thanks also to the wider DiSCOVERY study team: Ruth Mills, Kathryn Sams, Leanne Hague, Claire Duddy, Charlotte Wheeler, Maria Sanchez and Robert Kelly. Correspondence concerning this article should be addressed to Linda Birt, University of Leicester, United Kingdom.

## Data availability statement

The data that support the findings of this study are available on request from the corresponding author, [LB]. The data are not publicly available due to the sample size so full pseudonymized data sets may comprise the privacy of research participants.

## References

Amponsem, S., Wolverson, E., & Clarke, C., (2023). The meaning and experience of hope by people living with dementia as expressed through poetry. Dementia, 22(1), 125–143. 10.1177/14713012221137469

Bamford, C., Wheatley, A., Brunskill, G., et al. (2021). Key components of post-diagnostic support for people with dementia and their carers: A qualitative study. PloS one, 16(12). 10.1371/journal.pone.0260506

Birt, L., Poland, F., Csipke, E., & Charlesworth, G. (2017). Shifting dementia discourses from deficit to active citizenship. Sociology of Health & Illness, 39(2), 199–211. 10.1111/1467-9566.12530

Birt, L., Griffiths, R., Charlesworth, G., et al. (2020). Maintaining Social Connections in Dementia: A Qualitative Synthesis. Qualitative Health Research, 30(1), 23–42. 10.1177/1049732319874782

Birt, L., West, J., Poland, F., et al. (2023). Protocol for a realist evaluation of Recovery College dementia courses: understanding coproduction through ethnography. BMJ open, 13(12), e078248. 10.1136/bmjopen-2023-078248

Birt, L., Fox, C., Alam, J., et al. (2025). Co-produced, peer supported post-diagnostic dementia support: a realist evaluation of Recovery College courses. (In review)

Clarke, C. L., Wilcockson, J., Gibb, C. E., Keady, J., Wilkinson, H., & Luce, A. (2011). Reframing risk management in dementia care through collaborative learning. Health & Social Care in the Community, 19(1), 23–32. 10.1111/j.1365-2524.2010.00944.x

Department of Health. (2009). National Dementia Strategy Living well with dementia: A National Dementia Strategy Putting People First. https://assets.publishing.service.gov.uk/media/5a7a15a7ed915d6eaf153a36/dh_094051.pdf

Dooley, J., Webb, J., James, R., Davis, H., & Read, S. (2025). Exploring experiences of dementia post-diagnosis support and ideas for improving practice: A co-produced study. Dementia, 0(0). 10.1177/14713012241312845

Downs, M., & Lord, K. (2017). Person-Centered Dementia Care in the Community: A Perspective from the United Kingdom. Journal of Gerontological Nursing, 43(08), 11–17. 10.3928/00989134-20170515-01

Dufault, K., & Martocchio, B. C. (1985). Symposium on compassionate care and the dying experience. Hope: its spheres and dimensions. The Nursing clinics of North America, 20(2), 379–391.

Handley, M., Wheeler, C., Duddy, C., et al. (2024). Operationalising the Recovery College model with people living with dementia: a realist review. Aging & Mental Health, 28(8), 1078–1089. 10.1080/13607863.2024.2356878

Hayes, D., Hunter-Brown, H., Camacho, E. et al. (2023). Organisational and student characteristics, fidelity, funding models, and unit costs of Recovery Colleges in 28 countries: a cross-sectional survey. The Lancet Psychiatry, 10(10), 768–779. 10.1016/s2215-0366(23)00229-8

Hennelly, N., Cooney, A., Houghton, C., & O’Shea, E. (2021). Personhood and Dementia Care: A Qualitative Evidence Synthesis of the Perspectives of People With Dementia, The Gerontologist, 61(3), e85–e100. 10.1093/geront/gnz159

Hill, L., Roberts, G., Wildgoose, J., Perkins, R., & Hahn, S. (2010). Recovery and person-centred care in dementia: common purpose, common practice? Advances in Psychiatric Treatment, 16(4), 288–298. 10.1192/apt.bp.108.005504

Jackman, V., Wolverson, E., Clarke, C., & Quinn, C. (2024). A participatory approach to understand what might be most meaningful to people living with dementia in a positive psychology intervention. Aging & Mental Health, 28(8), 1090–1099. 10.1080/13607863.2023.2299967

Jethwa, J., Hunter-Brown, H., & Senkoy, A. Memory Services National Accreditation Programme (MSNAP) Standards: Standards for Memory Services. London; 2024. https://www.rcpsych.ac.uk/docs/default-source/improving-care/ccqi/quality-networks/memory-clinics-msnap/msnap-standards---9th-edition-(2024).pdf?sfvrsn=8c42ae6c_3

Jones-Hooker, C., & Tyndall, D. E. (2023). Application of case study research and ethnography methods: Lessons learned. Applied Nursing Research, 73, 151713. 10.1016/j.apnr.2023.151713

Jopling, K. (2017). AGE UK report: promising approaches to living well with dementia. https://www.ageuk.org.uk/siteassets/documents/reports-and-publications/reports-and-briefings/health--wellbeing/rb_feb2018_promising_approaches_to_living_well_with_dementia_report.pdf

Kenny, J., Asquith, I., Guss, R., et al. (2016). Facilitating an evolving service user involvement group for people with dementia: What can we learn? The Journal of Mental Health Training, Education and Practice, 11(2), 81–90. 10.1108/JMHTEP-09-2015-0046

Kitwood, T. (1997) Dementia reconsidered: The person comes first. Open University Press, Buckingham.

Kuske, S., Borgmann, S. O., Wolf, F., & Bleck, C. (2021). Emotional Safety in the Context of Dementia: A Multiperspective Qualitative Study. Journal of Alzheimer’s disease, 79(1), 355–375. 10.3233/JAD-201110

Leamy, M., Bird, V., Boutillier, C.L., Williams, J., & Slade, M. (2011). Conceptual framework for personal recovery in mental health: systematic review and narrative synthesis. British Journal of Psychiatry, 199(06), 445–452. 10.1192/bjp.bp.110.083733

Low, L.F., Swaffer, K., McGrath, M., & Brodaty, H. (2018). Do people with early stage dementia experience Prescribed Disengagement®? A systematic review of qualitative studies. International Psychogeriatrics, 30(6), 807–831. 10.1017/s1041610217001545

Lowen, C., Birt, L., & West, J. (2019). Recovery Colleges and dementia courses – a scoping survey, Mental Health and Social Inclusion, 23(4), 166–172. 10.1108/MHSI-08-2019-0024

Manzano, A. (2016). The craft of interviewing in realist evaluation. Evaluation, 22(3), 342–360. 10.1177/1356389016638615

Mason, B. (2022). Towards positions of safe uncertainty. Human Systems, 2(2), 54–63. 10.1177/26344041211063125

Miller, J.F. (1985). Inspiring Hope. The American Journal of Nursing, 85(1), 22. 10.2307/3463675

Miro (2022). Miro online whiteboard (no version provided). RealTimeBoard, Inc. www.miro.com

Moniz-Cook, E., & Mountain, G. (2023). The memory clinic and psychosocial intervention: Translating past promise into current practices. Frontiers in Rehabilitation Sciences, 4, 1052244. 10.3389/fresc.2023.1052244

Moniz-Cook, E., Manthorpe, J., Carr, I., Gibson, G., & Vernooij-Dassen, M. (2006). Facing the future: A qualitative study of older people referred to a memory clinic prior to assessment and diagnosis. Dementia. 5(3), 375–395. 10.1177/1471301206067113

Mukadam, N., & Livingston, G. (2012). Reducing the stigma associated with dementia: approaches and goals. Aging Health, 8(4), 377–386. 10.2217/ahe.12.42

NHS England (2024). Dementia programme and preparation for new Alzheimer’s disease modifying treatments. https://www.england.nhs.uk/long-read/dementia-programme-and-preparation-for-new-alzheimers-disease-modifying-treatments/https://www.england.nhs.uk/long-read/dementia-programme-and-preparation-for-new-alzheimers-disease-modifying-treatments/

Oh, H. (2013). The pedagogy of recovery colleges: clarifying theory. Mental Health Review Journal, 18(4). 10.1108/MHRJ-07-2013-0026

Palmer, V. J., Weavell, W., Callander, R., et al. (2019). The Participatory Zeitgeist: an explanatory theoretical model of change in an era of coproduction and codesign in healthcare improvement. Medical Humanities, 45(3), 247–257. 10.1136/medhum-2017-011398

Pawson, R., & Tilley, N. (1997). Realistic evaluation. Sage.

Perkins, R., Repper, J., Rinaldi. M., & Brown, H. (2012). Implementing Recovery through Organisational Change: Briefing paper. Centre for Mental Health. https://static1.squarespace.com/static/65e873c27971d37984653be0/t/668cedec6ea5392f6ac0ef31/1720511981544/1.Recovery-Colleges.pdf

QSR International Ltd. NVivo (released in March 2020). https://www.qsrinternational.com/nvivo-qualitative-data-analysissoftware/home

Reid, N., Khan, B., Soklaridis, S. et al. (2020). Mechanisms of change and participant outcomes in a Recovery Education Centre for individuals transitioning from homelessness: a qualitative evaluation. BMC Public Health, 20, 497. 10.1186/s12889-020-08614-8

Toney, R., Elton, D., Munday, E., et al. (2018). Mechanisms of Action and Outcomes for Students in Recovery Colleges. *Psychiatric services* (Washington, D.C.), 69(12), 1222–1229. 10.1176/appi.ps.201800283

van Corven, C. T. M., Bielderman, A., Wijnen, M., et al. (2021). Empowerment for people living with dementia: An integrative literature review. International Journal of Nursing Studies, 124, 104098. 10.1016/j.ijnurstu.2021.104098

Wall, S. S. (2015). Focused Ethnography: A Methodological Adaptation for Social Research in Emerging Contexts. Forum Qualitative Sozialforschung / Forum: Qualitative Social Research, 16(1). 10.17169/fqs-16.1.2182

Walsh, S., Merrick, R., Milne, R., Nurock, S., Richard, E., & Brayne, C. (2024). Considering challenges for the new Alzheimer’s drugs: Clinical, population, and health system perspectives. Alzheimer’s & Dementia, 20(9), 6639–6646. 10.1002/alz.14108

Watts, S., Cheston, R., Moniz-Cook, E., Burley, C., & Guss, R. (2014). Post-diagnostic support for people living with dementia. In R. Guss et al. Clinical psychology in the early-stage dementia care pathway. Leicester: British Psychological Society.

West, J., Birt, L., Wilson, D., Mathie, E., & Poland, F. (2022). A case study of co-production within a mental health Recovery College dementia course: Perspectives of a person with dementia, their family supporter and mental health staff. Frontiers in Rehabilitation Sciences, 3, 920496. 10.3389/fresc.2022.920496

Windle, G., Roberts, J., MacLeod, C., et al. (2023). ‘I have never bounced back’: resilience and living with dementia. Aging & Mental Health, 27(12), 2355–2367. 10.1080/13607863.2023.2196248

Wittenberg, R., Hu, B., Barraza-Araiza, L., & Rehill, A. (2019). Projections of older people living with dementia and costs of dementia care in the United Kingdom, 2019–2040. In CPEC Working Paper 5. https://www.infocoponline.es/pdf/Working-paper-5-Wittenberg-et-al-dementia.pdf

Wolverson, E.L., Clarke, C. and Moniz-Cook, E. (2010). Remaining hopeful in early-stage dementia: a qualitative study. Aging & Mental Health, 14(4), 450–460.

Wolverson, E., Hague, L., West, J., et al. (2024), Building an initial understanding of UK Recovery College dementia courses: a national survey of Recovery College and memory services staff, Working with Older People, 28(2), 108–119. 10.1108/WWOP-02-2023-0003

