## supplemental file 1 for "Role of peer-tutors with dementia in Recovery College dementia courses: an ethnographic account"

**Supplementary File 1: Recovery Colleges**

The first UK Recovery College was set up in 2009, growing to 85 by 2017 (Perkins, Meddings, Williams & Repper, 2018). The model has been replicated internationally with Recovery Colleges now in existence or developing in 22 countries (King & Meddings, 2019). The recovery model is flourishing in adult statutory mental health services, encouraged since 2011 by the Department of Health commissioned 'Implementing Recovery through Organisational Change' (ImROC) collaborative https://imroc.org/. A recovery-focused, peer-led psychoeducation approach is adopted and embedded within strategic care delivery in mental health Trusts, to complement clinical care and improve outcomes beyond a narrow focus on symptom reduction, to help people rebuild meaningful, satisfying lives, despite limitations caused by mental health difficulties (Perkins et al., 2018).

Recovery Colleges operate using five key linked conceptual processes - the CHIME recovery framework – robustly developed to underpin the term recovery in this context: **Connecting** with others, inspiring **Hope**, maintaining a positive **Identity**, finding **Meaning** and purpose in life outside of symptoms and **Empowering** control over life and a focus on strengths (Leamy, Bird, Boutillier, Williams & Slade, 2011). Recovery College courses offer access to distinctive peer support both from co-producing courses and/or attending them (Sommer, Gill & Stein-Parbury, 2018). Recovery Colleges create adult learning environments which moderate power dynamics between service users and staff, to reduce stigma and increase attendees' sense of hope and empowerment (Meddings, Byrne, Barnicoat, Campbell & Locks, 2014; Sommer et al., 2018; Zabel, Donegan, Lawrence & French, 2016). Attendees report developing novel coping strategies, improving self-worth, wellbeing and quality of life (Meddings et al., 2014; Rinaldi, Marland & Wybourn, 2012; Wilson, King & Russell, 2019; Zucchelli & Skinner, 2013). Implementing a recovery-focused approach through Recovery Colleges has been widely evidenced as bringing benefits as well as challenges to access (Allard et al., 2024; Bowness et al., 2023; Whitley et al., 2019).

A typical adult mental health Recovery College offers courses on mental health and recovery, designed to increase attendees' knowledge, skills and confidence in self-management of their own mental health and wellbeing. Courses range from one-off sessions to several sessions spread over a set number of weeks. All courses are co-produced and co-delivered by peer tutors - that is, people with lived/expert experience - and mental health staff. Peer tutors prepare for their role through having training to teach and support, and receive supervision to ensure any sensitive issues can be supported effectively. A theory of change model for Recovery Colleges has been co-developed within adult mental health contexts that identifies four mechanisms of change (Toney et al., 2018) empowering environment - opportunities for choices; shifting balance of power; enabling different relationships and connecting with peers; and facilitating personal growth through shared learning and strength-building. Outcomes include changes in the attendee including improved wellbeing, reinforced by life changes they could observe. This model is highly applicable to enabling desired outcomes for post-diagnostic support in dementia.

The CHIME Recovery framework (Leamy et al., 2011) as operationalised through Recovery Colleges, has clear links with the NICE-recommended person-centred care framework for dementia (Brooker & Latham, 2016), the Royal College of Psychiatrists Memory Services National Accreditation Programme (MSNAP; Jethwa, Fern, Abhayaratne & Wariabharaj, 2022) and the National Dementia Strategy objective to develop peer support and learning networks (Department of Health, 2009). Key domains for person-centred care are **valuing** people living with dementia and those (both informal family and friends and health and social care staff) who care for them; providing care that is **individualised**; understanding and acting from the **perspectives** of people living with dementia - which can reinforce connections, meanings and identities; and creating positive **social-psychological** environments - which can build hope and empowerment (Brooker & Latham, 2016).

*Five recovery processes giving the acronym CHIME*

**Connectedness** with others;

inspiring **Hope** and optimism about the future;

maintaining a positive **Identity**;

finding **Meaning** in life outside of symptoms;

and **Empowerment** with control over life and a focus on strengths. (Leamy et al., 2011)

**References**

Allard, J., Pollard, A., Laugharne, R., et al. (2024). Evaluating the impact of a UK Recovery College on mental well-being: pre- and post-intervention study. BJPsych open, 10(3), e87. https://doi.org/10.1192/bjo.2023.646

Bowness, B., Hayes, D., Stepanian, K., et al. (2023). Who uses Recovery Colleges? Casemix analysis of sociodemographic and clinical characteristics and representativeness of recovery college students. Psychiatric rehabilitation journal, 46(3), 211–215. https://doi.org/10.1037/prj0000532

Brooker, D., & Latham, I. (2016). *Person-centred dementia care making services better with the VIPS framework (2nd ed.)*. London Philadelphia: Jessica Kingsley Publishers.

Department of health. (2009). *National Dementia Strategy Living well with dementia: A National Dementia Strategy Putting People First*. https://assets.publishing.service.gov.uk/media/5a7a15a7ed915d6eaf153a36/dh_094051.pdf

Jethwa, J., Fern, M., Abhayaratne, C., & Wariabharaj, K. (2022). *Quality Standards for Memory Services Eighth Edition*. https://www.rcpsych.ac.uk/docs/default-source/improving-are/ccqi/quality- networks/memory-clinics-msnap/msnap-standards---8th-edition-(2022).pdf?sfvrsn=d8341549_2

King, T., & Meddings, S. (2019). Survey identifying commonality across international Recovery Colleges. *Mental Health and Social Inclusion,* https://doi.org/10.1108/mhsi-02-2019-0008

Leamy, M., Bird, V., Boutillier, C. L., Williams, J., & Slade, M. (2011). Conceptual framework for personal recovery in mental health: systematic review and narrative synthesis. *British Journal of Psychiatry*, 199(06), 445–452. https://doi.org/10.1192/bjp.bp.110.083733

Meddings, S., Byrne, D., Barnicoat, S., Campbell, E., & Locks, L. (2014). Co-delivered and co-produced: creating a recovery college in partnership. *The Journal of Mental Health Training, Education and Practice*, 9(1), 16–25. https://doi.org/10.1108/jmhtep-04-2013-0011

Perkins, R., Meddings, S., Williams, S., & Repper, J. (2018). *Recovery Colleges 10 Years On. ImROC.* https://static1.squarespace.com/static/65e873c27971d37984653be0/t/668c0c17506df25c98d01377/1720454168113/ImROC-Recovery-Colleges-10-Years-On.pdf

Rinaldi, M., Marland, M., & Wybourn, S. (2012). *Annual Report 2011 – 2012 South West London Recovery College*. http://rfact.org.au/wp-content/uploads/2015/05/SW-London-Recovery-College-evaluation-2011_12-v1-0.pdf

Sommer, J., Gill, K., & Stein-Parbury, J. (2018). Walking side-by-side: Recovery Colleges revolutionising mental health care. *Mental Health and Social Inclusion*, 22(1), 18–26.

https://doi.org/10.1108/mhsi-11-2017-0050

Toney, R., Elton, D., Munday, E., Hamill, K., Crowther, A., Meddings, S., Taylor, A., Henderson, C., Jennings, H., Waring, J., Pollock, K., Bates, P., & Slade, M. (2018). Mechanisms of Action and Outcomes for Students in Recovery Colleges. Psychiatric Services, 69(12), 1222–1229.

https://doi.org/10.1176/appi.ps.201800283

Wilson, C., King, M., & Russell, J. (2019). A mixed-methods evaluation of a Recovery College in South East Essex for people with mental health difficulties. Health & social care in the community, 27(5), 1353–1362. https://doi.org/10.1111/hsc.12774

Whitley, R., Shepherd, G., & Slade, M. (2019). Recovery Colleges as a mental health innovation. World Psychiatry, 18(2), 141-142. https://doi.org/10.1002/wps.20620

Zabel, E., Donegan, G., Lawrence, K., & French, P. (2016). Exploring the impact of the recovery academy: a qualitative study of Recovery College experiences. The Journal of Mental Health Training, Education and Practice, 11(3), 162–171. https://doi.org/10.1108/jmhtep-12-2015-0052

Zucchelli, F.A., & Skinner, S. (2013). Central and North West London NHS Foundation Trust's (CNWL) Recovery College: the story so far .... *Mental Health and Social Inclusion*, 17(4), 183–189. https://doi.org/10.1108/mhsi-07-2013-0023
