## supplementary file 2 for "Role of peer-tutors with dementia in Recovery College dementia courses: an ethnographic account"

**upplementary File 2:  Interview Topic Guides**

**<u>Topic guide 1</u> – peer tutors – people with dementia and family supporters**

Can you tell me about your experiences of co-producing and facilitating a Recovery College dementia course?

How did you become involved? Why did you become involved?

- *approached by a member of staff*
- *through the central Recovery College*
- *word of mouth or advert*

What have been the benefits of being involved?

What have been the challenges of being involved?

Can you tell me more about your role in co-developing the course?

- *who were you working with?*
- *how often did you meet and where*?

Was there already an idea for the course and how much do you think you helped to develop the course?

Were you happy with the level of involvement you had with developing the course?

Can you expand on that?

What support was provided for you as a coproduction partner?

- *financial*
- *practical (with technology/travel/accessing venues)*
- *emotional*

Who provided this support?

Did you experience any challenges for co-developing the course?

- *practical*
- *emotional*

- *time commitments*

How was your experience of working with the staff to co-produce the course?

- *supportive*
- *different from usual encounters with staff*
- *differences in power relations*

What do you see you gained from being involved with developing the course?

- *e.g. sense of purpose, new skills, confidence in skills, contributing by sharing own experience with others*

From your experience, do you think there are certain skills or motives that have helped you to co-produce the course?

- *confident with sharing experience*
- *willing to contribute*
- *ready to share experiences – can you expand on this?*

Can you tell me about your experience of running the course?

What support do you think you needed to run the course?

- *what worked well*
- *what did not work so well*

What do you think you have gained from running the course?

- *increase confidence*
- *chance to give back*

What do you think people gain from you running the course?

- *ideas that might help people live positively*
- *learning from and sharing experiences with others*

- *seeing someone with dementia leading the course and changing attitudes towards dementia*

- *increased knowledge of resources outside of the course (this could include links with other people)*

Are there negatives for you in running the course?

- *time-consuming*
- *difficult to talk about experiences?*
- *tiring*
- *level of expected involvement may not be in line with actual involvement*

Where and how is the course run?

- *online*
- *face-to-face*
- *what kind of venue?*
- *how often?*
- *for how long?*

Which do you prefer?

- *how do you think this impacts your experience of running the course?*
- *Why*

Which groups of people have you noticed come to the course?

- *people with dementia/supporters/staff?*
- *particular age group?*
- *background/ethnicity?*

Are there particular people/groups that you think aren't attending? Who? Why do you think this is?

- *transport/unable to access online*

- *lack of confidence*
- *aren't aware of the Recovery College*
- *think 'recovery' isn't relevant for people with dementia*
- *do not relate to the people running or attending course so think it is not for them*

How could more people be encouraged to attend dementia courses?

- *changes to how course is advertised (e.g. language/more widely advertised)*
- *buddy system/support with travel/technology*)

Are you involved with evaluating the course?

- *how is the course evaluated? (standard feedback form/questionnaire, more open-ended feedback welcomed?)*
- *is this information used to make changes to future courses?*
- *does it influence whether the course is run again?*

Anything else you would like to add?

**Topic guide 2 – staff tutors**

Can you tell me about your experiences of coproducing and facilitating a Recovery College dementia course?

How did you become involved? Why did you become involved?

What have been the benefits of being involved?

What have been the challenges?

**Setting up a course** What factors impact the decision to set up a Recovery College dementia course?

- *resources*
- *backing of certain individuals*
- *attitudes towards recovery in relation to dementia*
- *culture within the Trust*

What is the relationship between memory services and the Recovery College?

- *how do they complement one another?*
- *do they have differing aims?*
- *do memory services signpost people to the Recovery College?*

Who is the course open to?

- *people who have accessed services, supporters?*
- *staff?*
- *general public?*

How is the course advertised?

- *social media?*
- *through the Recovery College?*
- *through services?*
- *word of mouth?*
- *any other*

Are there particular groups of people you are aiming to reach with the Recovery College courses and what methods do you use to reach them?

Does this work?

**Coproduction** How important is coproduction to the course?

- *why is it important?*

What have been your experiences of coproducing the dementia course?

What would co-production of the course ideally look like?

- *is it possible? Why, or why not?*
  - *resources co-producers can access*
  - *time*

- *decision-making*

- What level of co-production do you consider good enough?

What have been the benefits for you in coproducing the course?

- *ability to work in line with values?*
- *help to re-engage with day-to-day job*
- *professional growth*
- *personally rewarding (how?)*
- *decreased stress/burnout?*

What are the challenges of coproduction?

- *embracing a different power dynamic/valuing each person's involvement? (For you? For the people you're coproducing with?)*
- *amount of time it takes*
- *any difficulties with working with people with dementia specifically? (If yes, what are they, how do you address and overcome these?)*

How do you find people to coproduce courses with?

- *people you've worked with clinically*
- *through the central Recovery College*
- *word of mouth*

Are there certain skills, characteristics, or experience that you look for in a coproduction partner?

- *confident with sharing their experience*
- *willingness to contribute*
- *emotionally 'robust' enough?*

Can you tell me about a time when you were struggling to work in a co-productive way and what you did to address this?

- *the fact that they might benefit from the process*

What support is provided for coproduction partners (people with dementia and supporters)?

- *financial?*
- *practical (with technology/travel/accessing venues)?*
- *psychological/emotional?*

Who provides this support?

Has coproducing the course impacted your day-to-day job?

- If yes, how and how have you managed this? *(Affected interactions with other people with dementia? More likely to apply recovery-focused principles?)*

**Running a course** Where/how is the course run?

- *online/face-to-face?*
- *what kind of venue?*

How do you think this impacts the delivery of the course?

- *are people with dementia less able to use technology to attend online?*
- *does the course being face-to-face encourage more discussion or building of relationships?*

What have been the positives of running the course?

- For you as a member of staff *(chance to gain holistic view of people with dementia, to see them recovering and thriving, increase personal wellbeing, changed attitudes towards recovery)*
- For peer tutors *(increase confidence, chance to give back)*
- For attendees (*empowerment and self-management, a safe space, learning from and sharing experiences with others, seeing peers leading and succeeding, reduced stigma and more positive sense of self, able to visualise living well, see self as more than diagnosis, ability to build relationships that last outside of the course)*

And the negatives? *(time-consuming alongside other responsibilities)*

Have you felt supported whilst running the course?

- *by the Trust (if applicable) at organisation lead level or manager level?*
- *by fellow coproducers?*
- *by other colleagues?*
- *with administrative staff?*

Who comes along to the courses?

- *people with dementia/supporters/healthcare staff?*
- *particular age group?*
- *background/culture/ethnicity?*

Are there particular people that you think aren't attending?

- Why do you think this is?
    - *transport/unable to access online*
    - *lack of confidence*
    - *aren't aware of the Recovery College*
    - *think 'recovery' isn't relevant for people with dementia*
    - *don't think the course is for them (why might that be?)*

How could more people be encouraged to attend dementia courses?

- *more accessible language*
- *more widely advertised*
- *buddy system/support with travel/technology*
- *ensuring adverts and materials 'speak' to the people trying to reach*

**Evaluating a course** Do you evaluate the course?

- How? *(feedback form/questionnaire, more open-ended feedback welcomed?)*

- Is this information used to make changes to future courses?
- Does it influence whether the course is run again?

Anything else you would like to add?

**<u>Topic guide 3</u> – attendees – people with dementia and family supporters**

Can you tell me about your experiences of attending a Recovery College dementia course?

How did you hear about the course?

- *from a member of staff / care team*
- *word of mouth*
- *social media*

Why did you decide to attend?

- *wanted to meet others in a similar situation*
- *wanted to learn strategies to live positively with dementia*
- *wanted to find out what a Recovery College is*

Where and how was the course run?

- *online/face-to-face?*
- *what kind of venue?*

Did you have any worries or concerns before joining the course?

- *getting there / joining online*
- *didn't feel confident enough, worried about talking or meeting others,*
- *what you might be expected to do during the course*
- *how you might feel during or after the course*
- *not sure if recovery is relevant to someone with dementia*

Do you think things could have been done to make the course easier to access?

- If yes, in what way?

How could more people be encouraged to attend Recovery College dementia courses?

- *easier to understand language*
- *more widely advertised*
- *easier ways to sign up*
- *buddy system / support with travel / technology*

Would you prefer courses online or in person?

- Why?

What have been the positive outcomes from attending the course?

- *learning from and sharing experiences with others,*
- *seeing people with dementia running the course*
- *improving how I see myself and my situation*
- *knowing where I might get help or thinking about how I can better manage*

What have been the negative points from attending the course?

How do you think the course could have been improved?

- *ratio between people with dementia / family supporters and staff*
- *did everyone get the chance to speak if they wanted to?*
- *did you see it as a safe space?*

Anything else you would like to add?

**Topic guide 4 – attendees – staff**

Can you tell me about your experiences of attending a Recovery College dementia course?

How did you hear about the course?

- *from another member of staff*

- *word of mouth*
- *social media*
- *through central Recovery College*
- *previous attendance at Recovery College*

Why did you decide to attend?

- *thought learning would be helpful in day-to-day role*
- *wanted to learn more about recovery and dementia*
- *wanted a different way of working with people with dementia*
- *wanted to find out more about the Recovery College*

Were there any barriers to you attending?

- *taking time out of work*
- *permission from line manager*

Or how were you encouraged to attend and why?

- *recommended by colleagues*
- *part of CPD*
- *considering role of coproducer*

Where and how was the course run?

- *online*
- *face-to-face? What kind of venue?*

Did this work for you? Why or why not?

Do you think the course could have been more accessible?

- If yes, in what way?

How could more people be encouraged to attend Recovery College dementia courses?

- *more accessible language (less recovery-focused?)*
- *more widely advertised*
- *support with travel/technology*

What have you gained from attending the course?

- *learning from and sharing experiences with others*
- *seeing people with dementia leading, succeeding, and living positively*
- *reduced stigma and changed attitudes towards people with dementia*

Were your expectations for the course met?

- *what were they?*
- *how and why or why not?*

How do you think the course could have been improved?

- *ratio between lived and professional experience*
- *how the course was facilitated*
    - *did everyone get the chance to speak if they wanted to*
    - *did it feel like a safe space?*

What was your understanding of recovery in the context of dementia before attending? Did this change after attending the course?

- *not recognised as important by colleagues*

Was there any learning that you will take back to your day-to-day role?

Anything you would like to add?

**<u>Topic Guide 5</u> – Relating to Equality Diversity and Inclusion**

This part of the xx study is about trying to understand what we need to consider in Recovery College dementia courses to best support people from ethnic and cultural minorities with understanding dementia. So this is a very open, exploratory interview – not seeking a particular angle or answer at all, just really interested in hearing your views.

In our prior conversation [*prep for interview*], you mentioned that people from ethnic and cultural minority groups are under-represented in both your memory services and the Recovery College course. So thinking about the whole process and pathway, why do you feel that may be the case? What do you feel is the 'sticking point'?

*[You can think about it from the angle of the service users themselves, or other healthcare professionals, or services, systems]*

To what extent do you feel that people may feel excluded from services or are choosing not to engage?

Where do you feel that people seek or receive a diagnosis and post-diagnostic support if not from formal health services? Do you know of any other services which pick people up later in their journey?

What do you feel could be some of the unmet needs that ethnic minority people with dementia could have post-diagnosis due to current levels of support, if any?

What do you feel could be some of the wider drivers of under-representation in this group the Recovery College? i.e. geography, socioeconomic, health beliefs of dementia etc

**Moving Ahead:**

1. What do you feel we in Recovery Colleges and/or the wider health service could do better to attract ethnic and cultural minorities into courses?

2. What do you feel that Recovery Colleges could offer ethnic minority participants that is different or unique to other forms of support?

3. Does your Recovery College currently support or pay for the additional costs of running the courses i.e. travel costs, room, refreshments etc? How is this paid for? Is there anything else that you would like funding for to engage minorities in the course that may be helpful?

4. How do you feel that your current dementia Recovery College courses could or would need to change to better engage and support ethnic and cultural minority people?

ENDS
